# The effect of COVID-19 on critical care research during the first year of the pandemic: A prospective longitudinal multinational survey

**DOI:** 10.1101/2020.10.21.20216945

**Authors:** Mark Duffett, Deborah J Cook, Geoff Strong, Jan Hau Lee, Michelle E Kho

**Author notes:** **Corresponding author**: Mark Duffett, McMaster Children’s Hospital, 1200 Main St. West, Room 1E1A, Hamilton, ON L8N 3Z5.

## Abstract

**Importance:** The COVID-19 pandemic has increased the need for high-quality evidence in critical care, while also increasing the barriers to conducting the research needed to produce such evidence.

**Objective:** To determine the effect of the first wave of the COVID-19 pandemic on critical care clinical research.

**Design:** Monthly electronic survey (March 2020 - February 2021).

**Setting:** Adult or pediatric intensive care units (ICUs) from any country participating in at least one research study before the COVID-19 pandemic.

**Participants:** We recruited one researcher or research coordinator per center, identified via established research networks.

**Intervention(s):** None

**Main Outcome(s) and Measure(s):** Primary: Suspending recruitment in clinical research; Secondary: impact of specific factors on research conduct (5-point scales from no effect to very large effect). We assessed the association between research continuity and month, presence of hospitalized patients with COVID-19, and population (pediatric vs. adult ICU) using mixed-effects logistic regression.

**Results:** 127 centers (57% pediatric) from 23 countries participated. 95 (75%) of centers suspended recruitment in at least some studies and 37 (29%) suspended recruitment in all studies on at least one month. The proportion of centers reporting recruitment in all studies increased over time (OR per month 1.3, 95% CI 1.2 to 1.4, p < 0.001), controlling for hospitalized patients with COVID-19 and type of ICU (pediatric vs. other). The five factors most frequently identified as having a large or very large effect on clinical research were: local prioritization of COVID-19 specific research (68, 54%), infection control policies limiting access to patients with COVID-19 (61, 49%), infection control policies limiting access to the ICU (52, 41.6%), increased workload of clinical staff (38, 30%), and safety concerns of research staff (36, 29%).

**Conclusions and Relevance:** Decisions to pause or pursue clinical research varied across centers. Research activity increased over time, despite the presence of hospitalized patients with COVID-19. Guiding principles with local adaptation to safely sustain research during this and future pandemics are urgently needed.

**Key Points:** *Question:* What was the effect of the COVID-19 pandemic on research in 127 adult and pediatric intensive care units (ICUs) between March 2020 and February 2021?

*Findings:* 95 (75%) centers suspended recruitment into at least some studies. Active recruitment into studies increased over time (OR per month 1.3, 95% CI 1.2 to 1.4, p < 0.001), controlling for ICU type and the presence of patients with COVID-19.

*Meaning:* Research activity varied across centers and increased over time, despite the presence of hospitalized patients with COVID-19. Guiding principles to safely sustain research during this and future pandemics are urgently needed.

## Introduction

The coronavirus disease 2019 (COVID-19) pandemic demands a rapid, coordinated critical care clinical and research response, while also presenting barriers to conducting such research. Infection control precautions to reduce viral spread such as conserving personal protective equipment (PPE) and minimizing the number of personnel in hospitals have challenged the capacity to conduct clinical research in the face of the pandemic. Decisions to pause or pursue research may depend on the severity of the pandemic in a given jurisdiction and may vary over time as it advances or abates.

Both patients with and without COVID-19 may be eligible to participate in research studies during the pandemic. Focusing solely on COVID-19-specific research risks disadvantaging the patients without COVID-19 who may benefit from research. To better understand how intensive care units (ICUs) worldwide have attempted to balance such competing mandates, our objectives were to describe the effect of the first year of the pandemic. We studied the extent to which study recruitment stopped in critical care studies, and factors influencing the continuation of this research.

## Methods

We conducted a monthly online survey administered from March 8, 2020 to February 28, 2021, approaching representatives from adult and pediatric ICUs if they were enrolling patients in at least one clinical research study before the World Health Organization declaration of the pandemic. We recruited sites through research networks (Canadian Critical Care Trials Group, Pediatric Acute Lung Injury and Sepsis Investigators, Australia and New Zealand Intensive Care Society Clinical Trials Group, Pediatric Acute and Critical Care Medicine Asian Network), and Twitter. After Research Ethics Board (REB) approval we sent monthly surveys to one investigator or research coordinator at each site with up to two reminders to non-respondents each month. We included 33 items that captured site demographics, presence of patients with COVID-19 in the hospital and the ICU, characteristics of any clinical research studies underway or stopped. For the first 6 months we also included questions about perceived barriers to research conduct. The survey instrument was in English. No incentive was provided. Survey completion implied informed consent.

For analysis, children’s hospitals housed within larger hospitals and each site of multi-site hospitals were considered separate centers. For centers with multiple ICUs, each was considered separately or as one center, per local self-report. We retained all complete and partial surveys, using the number of responses for each question as the denominator. We reported data as median (interquartile range [IQR]) or count (percentage). We used Chi-square tests to compare the characteristics of centers continuing recruitment in all studies versus those that did not. We used mixed effects logistic regression to examine the association between time (months since the start of the pandemic) and research continuity, controlling for center type (exclusively pediatric vs other) and the presence of patients with COVID-19 in that hospital on that month. We report odds ratios (OR) with their corresponding 95% confidence intervals (CI) and used p <0.05 as the criterion for statistical significance. We used R version 3.6.3 (R Foundation for Statistical Computing, Vienna, Austria) to perform the analysis.

## Results

ICUs from 127 centers in 23 countries participated: 104 (82%) academic and 72 (57%) exclusively pediatric (Figure 1). Countries with the highest representation were Canada (37, 29%), United States (30, 24%), and Australia (21, 17%). The number of participating centers each month varied from 60 to 107, with 60 (47%) contributing data for at least 10 of the 12 months. Overall, 116 (91%) reported patients with COVID-19 in their hospital and 85 (67%) reported patients with COVID-19 in their ICU on at least one month; Figure 2 shows the monthly prevalence.

**Figure 1:**
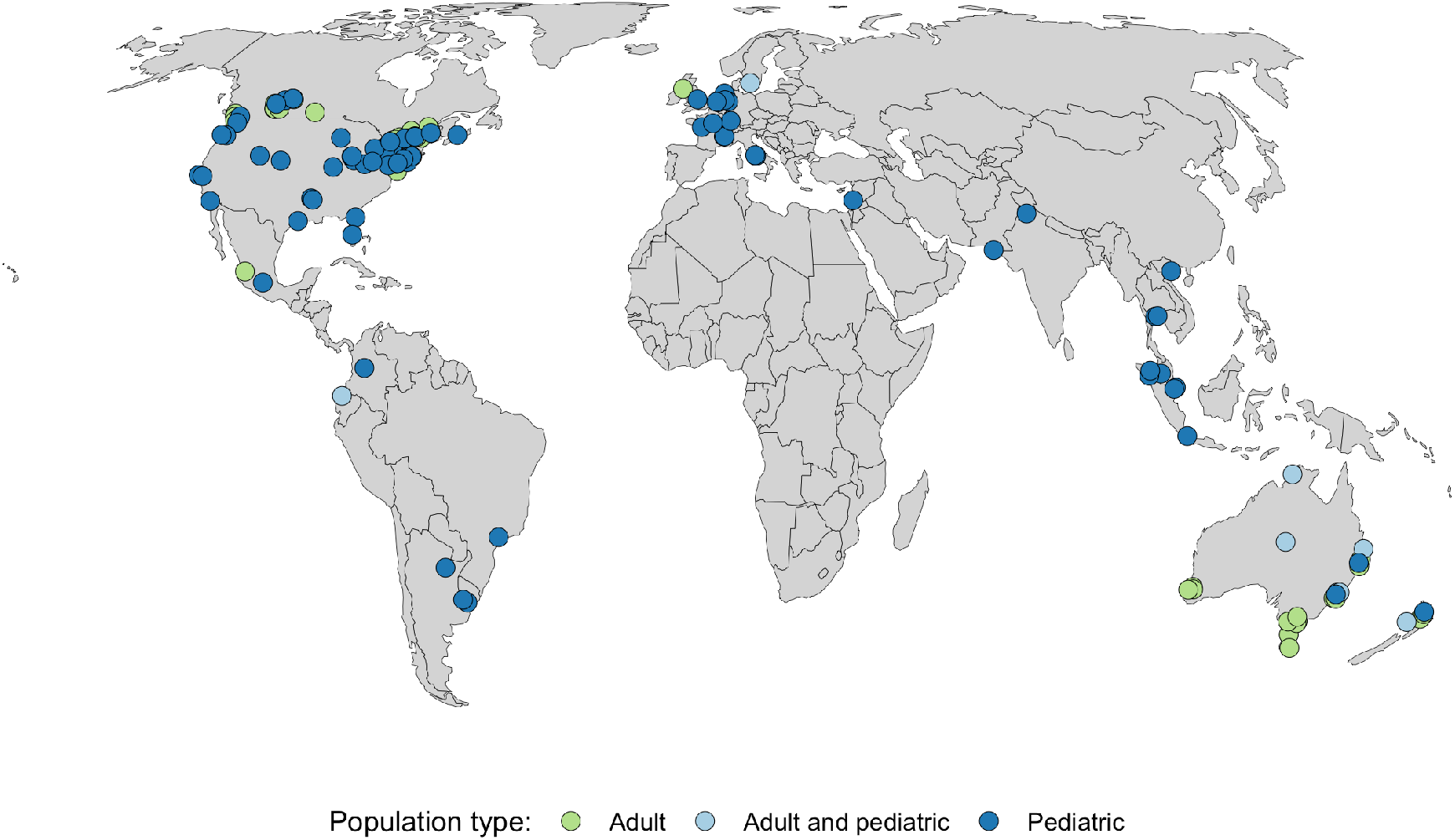
Participating centers.

**Figure 2:**
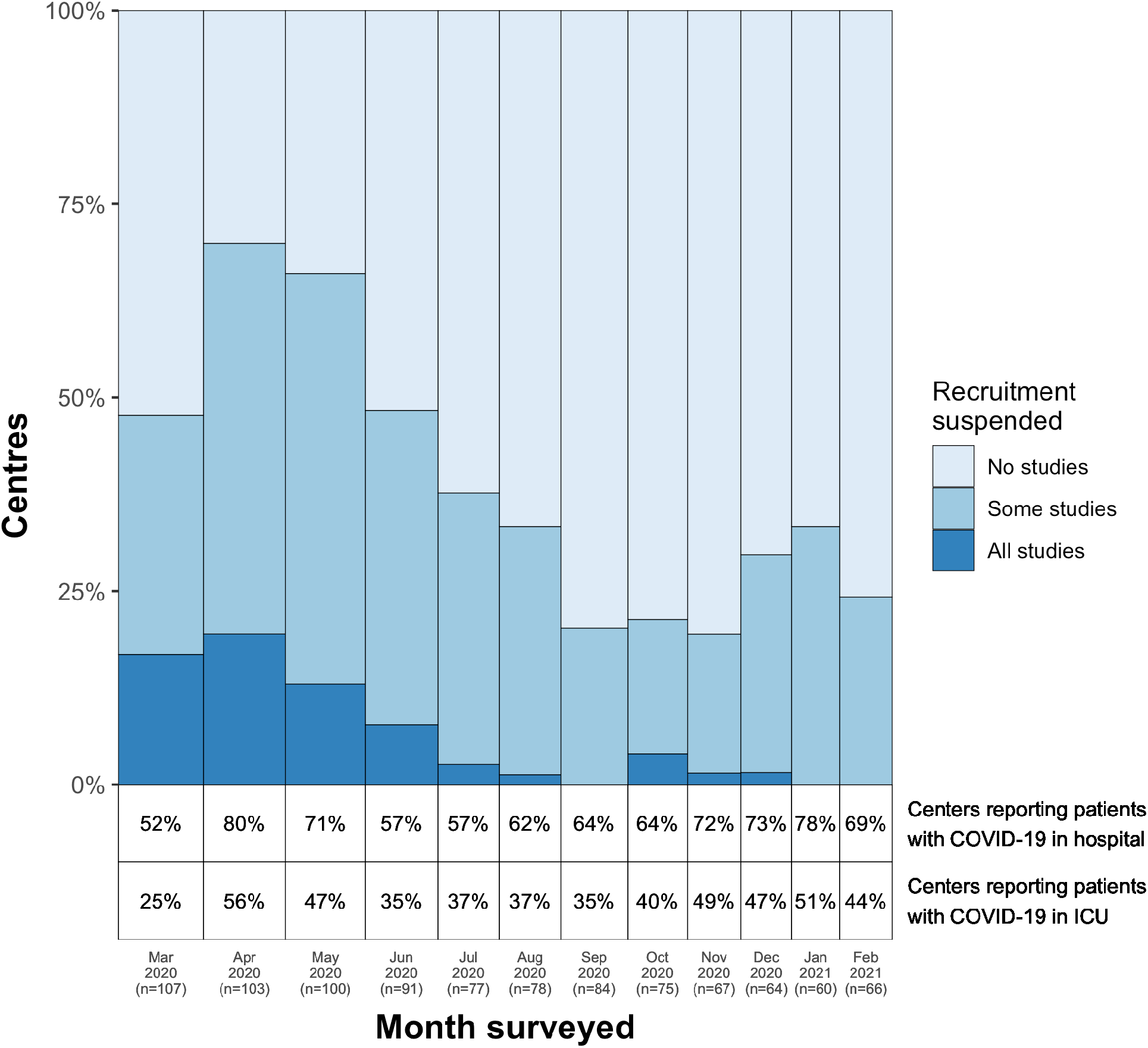
Centers suspending recruitment in clinical research studies due to COVID-19 pandemic.

In the first 6 months, 105 (83%) respondents reported preparing for COVID-19-specific studies, and 112 (89%) reported enrolling patients in COVID-19 specific studies, which varied from 30% in March to 84% in August. The percentage of centers reporting that their REB expedited COVID-19 specific research increased from 58% in March to 88% in August (p for trend <0.001).

On at least one month, 95 (75%) of centers suspended recruitment in some studies and 37 (29%) suspended recruitment in all studies. Pediatric centers were not significantly more likely than adult centres to suspend recruitment in all studies at some point (OR 1.6, 95% CI 0.8, 3.7, p = 0.30). Centres with hospitalized patients with COVID-19 were also not significantly more likely to suspend recruitment in all studies at some point (OR 0.9, 95% CI 0.2, 4.8, p>0.99).

Overall, 31 (25%) centers continued recruitment in all studies for the whole study period. The percentage of centers suspending recruitment varied by month, with 15% to 58% suspending recruitment in some studies and 0% to 18% suspending recruitment in all studies. (Figure 2). Active recruitment in all studies increased over time (OR per month 1.3, 95% CI 1.2 to 1.4, p < 0.001), controlling for hospitalized patients with COVID-19 and type of ICU (pediatric vs. other). The subgroup of centres that submitted at least 10 of the 12 surveys showed a similar pattern (Figure 2a, Appendix), suggesting that differential loss to follow-up was not a major contributor to the changes over time.

Figure 3 shows the reported effect of specific factors on research conduct. The five factors most frequently identified as having a large or very large effect on research conduct in any month were: the local prioritization of COVID-19 specific research (54%), infection control policies limiting access to patients with COVID-19 (49%), infection control policies limiting access to the ICU (42%), increased workload of clinical staff (30%), and safety concerns of research staff (29%).

**Figure 3:**
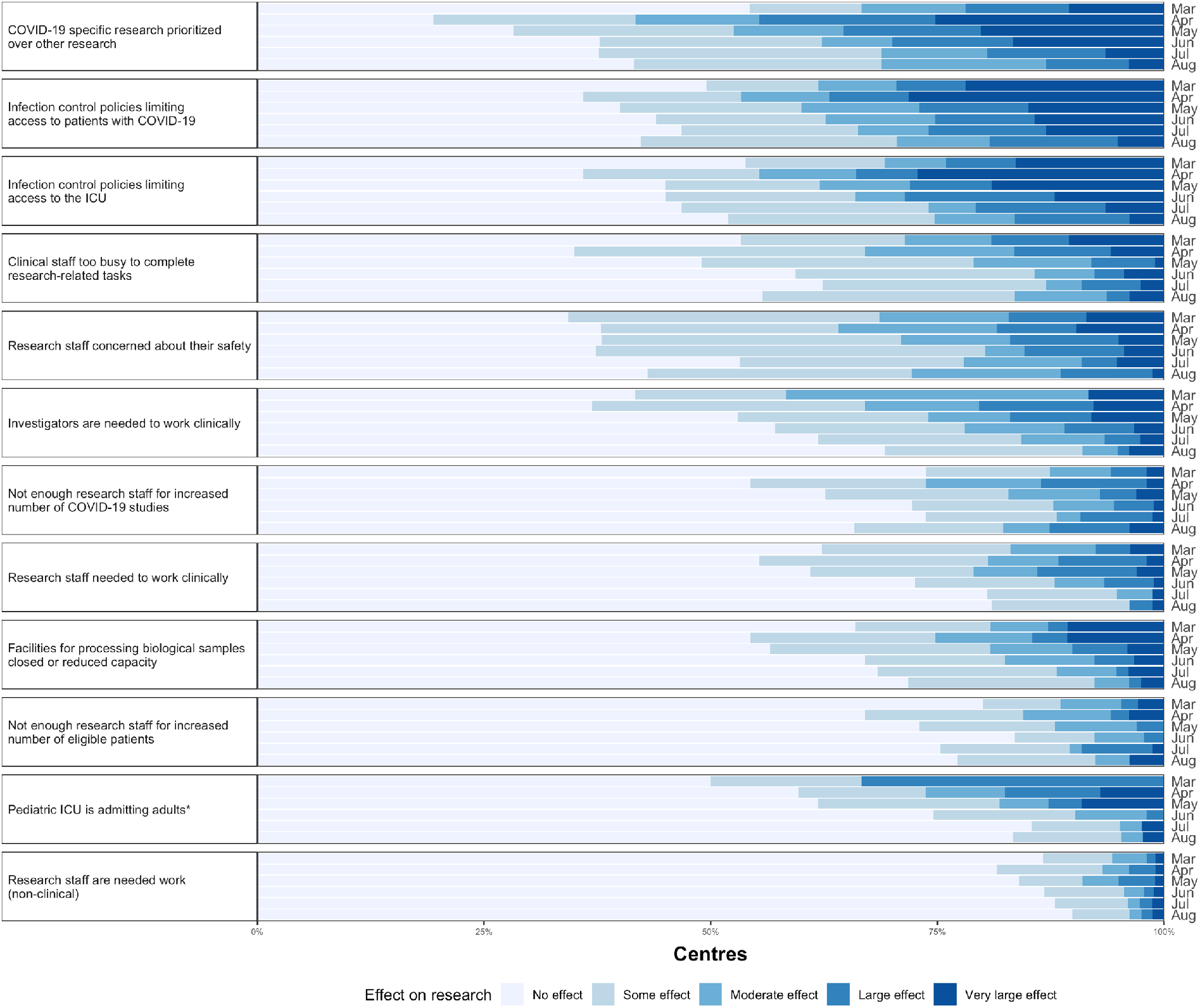
Effects of specific factors on critical care research. *Only pediatric centers included in the denominator for this question. These questions were included in the survey for the first 6 months (March – August 2020). The median (IQR) number of responses for each question per month was 91 (77, 103).

## Discussion

In this monthly survey of 127 centers in 23 countries we observed that decisions to suspend recruitment in clinical research during the first wave of the pandemic varied across centers and decreased over time, despite the presence of hospitalized patients with COVID-19. These substantial effects, even in countries such as Canada and Australia that were not overwhelmed in the first pandemic wave, likely underestimate the true effect on research activity globally. Despite fewer children with COVID-19 admitted to hospital and even fewer admitted to ICUs, PICUs reported being just as likely to experience clinical research disruptions as adult ICUs. One potential explanation for this is the need for surge capacity planning that required pediatric ICUs to admit adult patients with COVID-19 (Figure 3).^1,2^ This is particularly evident in the early period of our survey (March – May 2020).

Amidst the challenges to conducting research during the pandemic, several much-needed large multi-center RCTs of therapies for COVID-19 have been launched and completed.^3–5^ Learning ^6,7^ from these studies, investigators around the world can adapt research logistics for COVID-specific and non-COVID-specific studies, to best suit the local context to ensure timely continuation and completion of pertinent clinical research studies.^8,9^

Strengths of this study include prospective, monthly data collection during the first wave of the pandemic and representation of both adult and pediatric ICUs. Limitations include the convenience sample of participating centers and limited representation from regions with the highest demand for critical care resources during the first wave of the pandemic.

## Conclusions

Decisions to pause or pursue clinical research varied across centers in the first wave of the pandemic. Research activity increased over time, despite the presence of hospitalized patients with COVID-19. Guiding principles and local adaptations to safely sustain research in future months are urgently needed.

## Data Availability

Data for this manuscript is available from the authors.

## Appendix

**Figure 2a:**
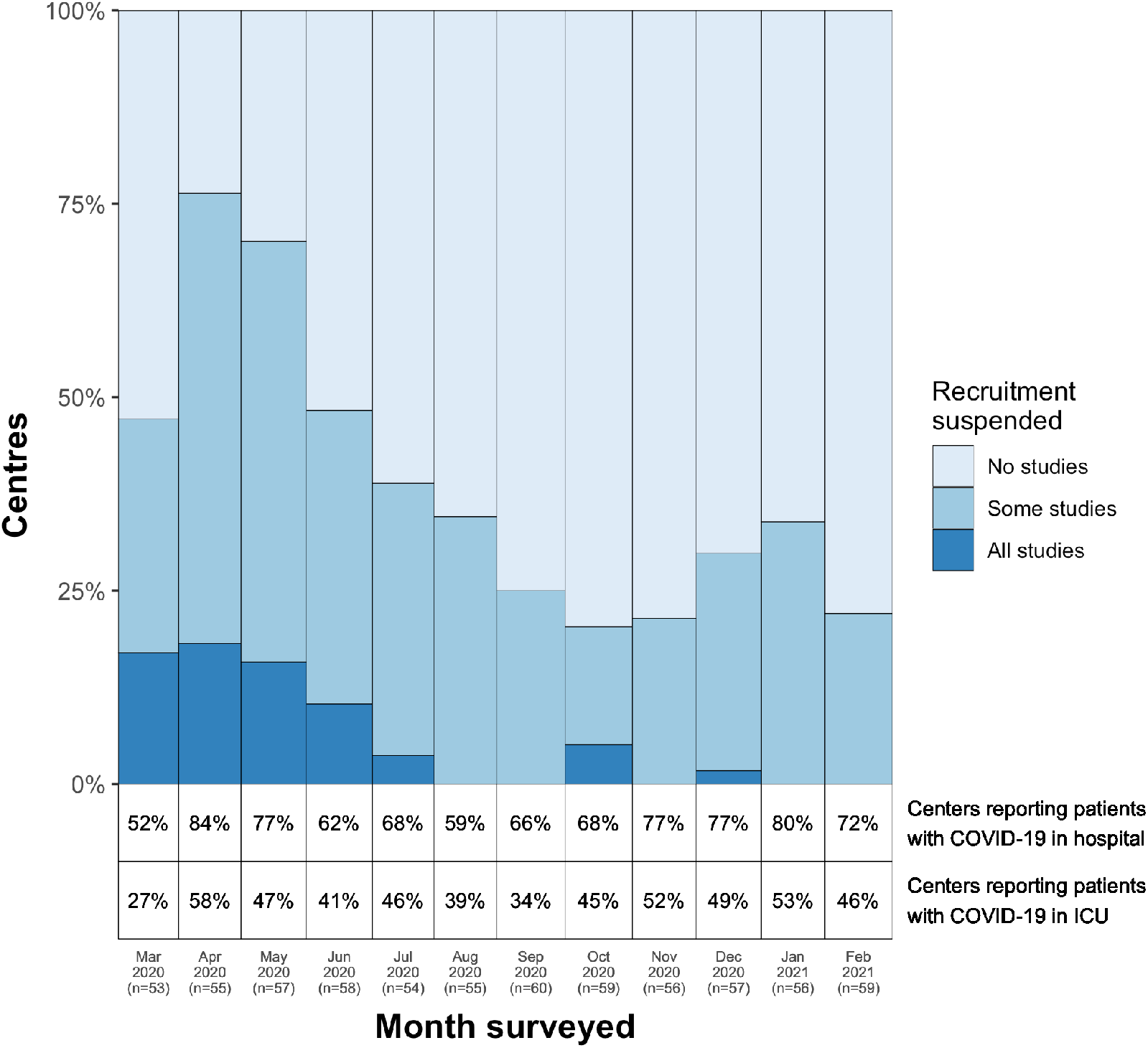
Centers suspending recruitment in clinical research studies due to COVID-19 pandemic (centres submitting surveys for at least 10 of the 12 months)

